# Automatic X-ray COVID-19 Lung Image Classification System based on Multi-Level Thresholding and Support Vector Machine

**DOI:** 10.1101/2020.03.30.20047787

**Authors:** Lamia Nabil Mahdy, Kadry Ali Ezzat, Haytham H. Elmousalami, Hassan Aboul Ella, Aboul Ella Hassanien

**Author notes:** Scientific Research Group in Egypt (SRGE), http://www.egyptscience.net.

## Abstract

The early detection of SARS-CoV-2, the causative agent of (COVID-19) is now a critical task for the clinical practitioners. The COVID-19 spread is announced as pandemic outbreak between people worldwide by WHO since 11/ March/ 2020. In this consequence, it is top critical priority to become aware of the infected people so that prevention procedures can be processed to minimize the COVID-19 spread and to begin early medical health care of those infected persons. In this paper, the deep studying based totally methodology is usually recommended for the detection of COVID-19 infected patients using X-ray images. The help vector gadget classifies the corona affected X-ray images from others through usage of the deep features. The technique is useful for the clinical practitioners for early detection of COVID-19 infected patients. The suggested system of multi-level thresholding plus SVM presented high accuracy in classification of the infected lung with Covid-19. All images were of the same size and stored in JPEG format with 512 * 512 pixels. The average sensitivity, specificity, and accuracy of the lung classification using the proposed model results were 95.76%, 99.7%, and 97.48%, respectively.

## 1. Introduction

The confirmed SARS-Cov-2, the causative agent of Wuhan new corona virus disease 2019 (COVID-19) cases exceeded SARS-CoV-1, the causative agent of severe acute respiratory syndrome (SARS) cases. At the time of writing this paper, there are currently +740,201 confirmed cases +28,000 critical/serious cases and +35,026 deaths worldwide from the COVID-19 pandemic outbreak as of March 30, 2020, 11:23 GMT. Coronaviruse is often presented with a novel word, as a new strain can be within the virus family, we have all discovered beforehand. As indicated by the World Health Organization (WHO), Coronaviruses belong to a large family that varies from cold to serious disease [1,8]. These diseases can affect humans and animals. The strain that began to spread in Wuhan, the capital of China’s Hubei region, was identified from two completely different Coronaviruses namely, Severe acute respiratory syndrome (SARS) and the Middle east respiratory syndrome (MERS). Symptoms of coronary virus infection increase the severity of respiratory complications such as respiratory disorder, kidney disorder and fluid development in the lungs.

By comparison, SARS killed 774 people in 2003 mostly in China, the epicenter of the pandemic outbreak. COVID-19 and SARS are spread across continents, infecting animals and humans, and using similar mechanisms to enter and injure the cell. On the front line, the tactical response to COVID-19 is similar to that of the SARS 2003 outbreak however, there is one major difference, the COVID-19 emergency is occurring in a much more digitized and connected to our world. The amount of data produced from the dawn of humankind through 2003 is generated today within a few minutes. Furthermore, advanced intelligence models, such as those based on artificial intelligence (AI) and machine learning (ML), have shown great potential in tracing the source or predicting the future spread of infectious diseases. It is therefore imperative to leverage big data and intelligent analytics and put them to good use for public health.

A broad range of means of COVID-19 medical diagnosis can be conducted to detect the confirmed cases of COVID-19 [8]. These means are contributing together to daily count up each new confirmed case using electronic health records (EHR). These means of COVID-19 medical diagnosis include clinical characteristics and radiologist’s diagnosis. Clinical characteristics involves human temperature monitoring and reverse transcription polymerase chain reaction (RT-PCR) [9]. Based on the human body temperature, patients’ body temperatures range within ranges of 36·5–38·8°C [11]. Accordingly, available thermal sensors or thermal images can be collected as instantaneous data to investigate the potential COVID-19 [8]. Reverse transcription polymerase chain reaction (RT-PCR) is a laboratory technique that is used for testing COVID-19 using a blood sample of the case. Using real time RT-PCR test confirms the actual number of confirmed cases globally [13,14]. Moreover, point-of-care testing, Immunoglobulin M (IgM) and detection of antibodies can be applied to detecting infected cases [6,16].

The gold standard test to detect the COVID-19 confirmed cases is quantative reverse transcriptase PCR (qRT-PCR). However, this approach needs a CDC guide’s sample collection, qualified microbiology expertise, time from 4hrs up to 6hrs. Therefore, medical imaging is very important candidate in screening for the COVID-19 cases [17]. Radiologist’s diagnosis involves computed tomography (CT) scans, chest X-ray (CXR) radiographs [10]. COVID-19 symptoms can be effectively detected using CT or X-ray images. Based on the chest CT scans During recovery, radiologists can detect the (COVID-19) pneumonia and the stage of patient recovery or deterioration. Automated artificial intelligence models can accurately provide early detection for the diagnosis of the cases of COVID-19 by detecting the early lung damage signs in the images. An inception artificial neural networks (ANNs) have been applied for binary classification for infected with COVID-19 or health persons using 1,119 CT images [18]. The modified the Inception transfer-learning model produced accuracy of 89.5% with 0.88 and 0.87 for specificity and sensitivity, respectively using internal validation dataset. On the other hand, model produced accuracy of 79.3% with 0.83 and 0.67for specificity and sensitivity, respectively using external validation dataset. Therefore, the findings demonstrate the reliability of deep learning to find COVID-19 radiological features based on terms of time and accuracy.

U-Net++ neural network model has been developed for processing over 6000 CT scans to classify the cases of patients into COVID-19 infected or not infected [19]. The model produced 93.55% and 100% for specificity and sensitivity, respectively. In addition, the model presented negative positive value (NPV) of 100%, positive value (PPV) of 84.62%, and a total accuracy of 95.24%. The model greatly helps radiologists by decreasing the reading time of CT scans by 65% [19]. On the other hand, 3-category classification model is designed to distinguish the COVID-19 cases where both Song et al. (2020) and Xu et al. (2020) have applied feature extraction using Feature Pyramid Network and several fully-connected layers for cases classifications [20, 21]. The feature extraction using Feature Pyramid Network model produces an acceptable total accuracy of 86.7% using CT scans. The fully-connected layers model can accurately detect the COVID-19 cases with AUC of 0.99 and sensitivity of 0.93.

As a result of this surveying, Although the diagnosis of COVID-19 is based firstly and mainly on the presence of clinical signs related to respiratory systems and more specifically an pneumonia related signs (e.g., dry cough, fatigue, myalgia, fever, and dyspnea) as well as history of recent (two weeks apart) exposure to a known confirmed cases, a broad range of means of COVID-19 rapid bed side, in field or point of care (POCT) medical diagnostics to help in this pandemic outbreak screening already have been developed in the few past weeks or still under developing for few upcoming days such as IgM/IgG based lateral flow (lateral immunochromatographic assay), Nucleic acid lateral flow (NALF), CRISPR-cas13 lateral flow (CASLFA) and radiological imaging. Although, CT scans and X-ray imaging are time consuming and exhaustive even for expert radiologists, it is characterized by that the highest sensitivity diagnostic method in comparison to the gold standard (qRT-PCR) and other already developed or will be developed rapid diagnostics. So, relying on radiological imaging in screening and diagnosis of COVID-19 is very medically meaningful and therefore a need to automated system based on Artificial Intelligence (AI) tools could be accurately provide automated, less exhaustive for medical imaging workers, early detection, follow up method of COVID-19 cases and at same time method of differentiation of the radiological images of affected lung due to COVID-19 from other causes of lung affections especially MERS which caused by (MERS-CoV) and SARS which caused by (SARS-CoV-1) which are another viruses belonging to *Coronavirdeae* viruses family, the same family to which the COVID-19 causative agent (SARS-CoV-2) is belonging and all three viruses have an overlapping clinical signs and affected lung radiological image patterns but a differences and uniqueness still present.

A radiological imaging of confirmed COVID-19 infected patient typically shows either patchy or diffuse asymmetric airspace opacities, similar to the SARS and MERS associating pneumonia. A bilateral lung affection at first few days of infection in 98% of cases was reported at the first report of patients with COVID-19, with two different patterns according to intensive care unit (ICU) association or not and it is described as follow, patients inside intensive care units (ICUs) showed consolidation pattern while patients outside (ICUs) showed ground-glass predominant pattern. Also a 21 confirmed COVID-19 cases investigatory study showed chest radiological images with lung affections in 86% of patients, with a majority of 89% having bilateral lung affections [28]. Peripheral lung affection with consolidation or multifocal ground-glass predominant opacities was reported in 29% and 57% of cases, respectively. As well, a COVID-19 confirmed seven people family cluster chest radiological images showed bilateral patchy ground-glass predominant opacities with greater involvement of older family member’s lung than those young members [27,29]. Although the imaging features closely similar and overlapping those associating of SARS and MERS, the bilateral lungs involvement on initial imaging is more likely to be seen with COVID-19; as those associating SARS and MERS are more predominantly unilateral.

Through detecting and analyzing the previously mentioned early lung affection patterns present in the radiological images. This paper presents automatic x-ray COVID-19 lung image classification system based on multi-level thresholding and supported vector machine. The study is usually recommended for the early detection of COVID-19 infected patients using X-ray images. The help vector gadget classifies the COVID-19 affected lung X-ray images from others through usage of the deep features. The technique is useful for the clinical practitioners for early detection of COVID-19 infected patients. The suggested system version, multi-level thresholding followed by applying SVM to classify whether the people are infected or uninfected with COVID-19.

## 2 Materials and Methods

### 2.1 Dataset Characteristics

In total, 40 contrast-enhanced lungs X-ray with 512×512 in-plane resolution. 15 normal lung images are from the Montgomery County X-ray Set, while other 25 images are infected lungs with COVID-19 from covid-chestxray-dataset-master. In total, 20 lungs X-ray with 512×512 in-plane resolution. Among them, 10 normal lung images are from the Montgomery County X-ray Set, and 10 are from the public database covid-chestxray-dataset-master [1]. Figure (1) shows sample of the images we have used in this study.

**Fig 1.**
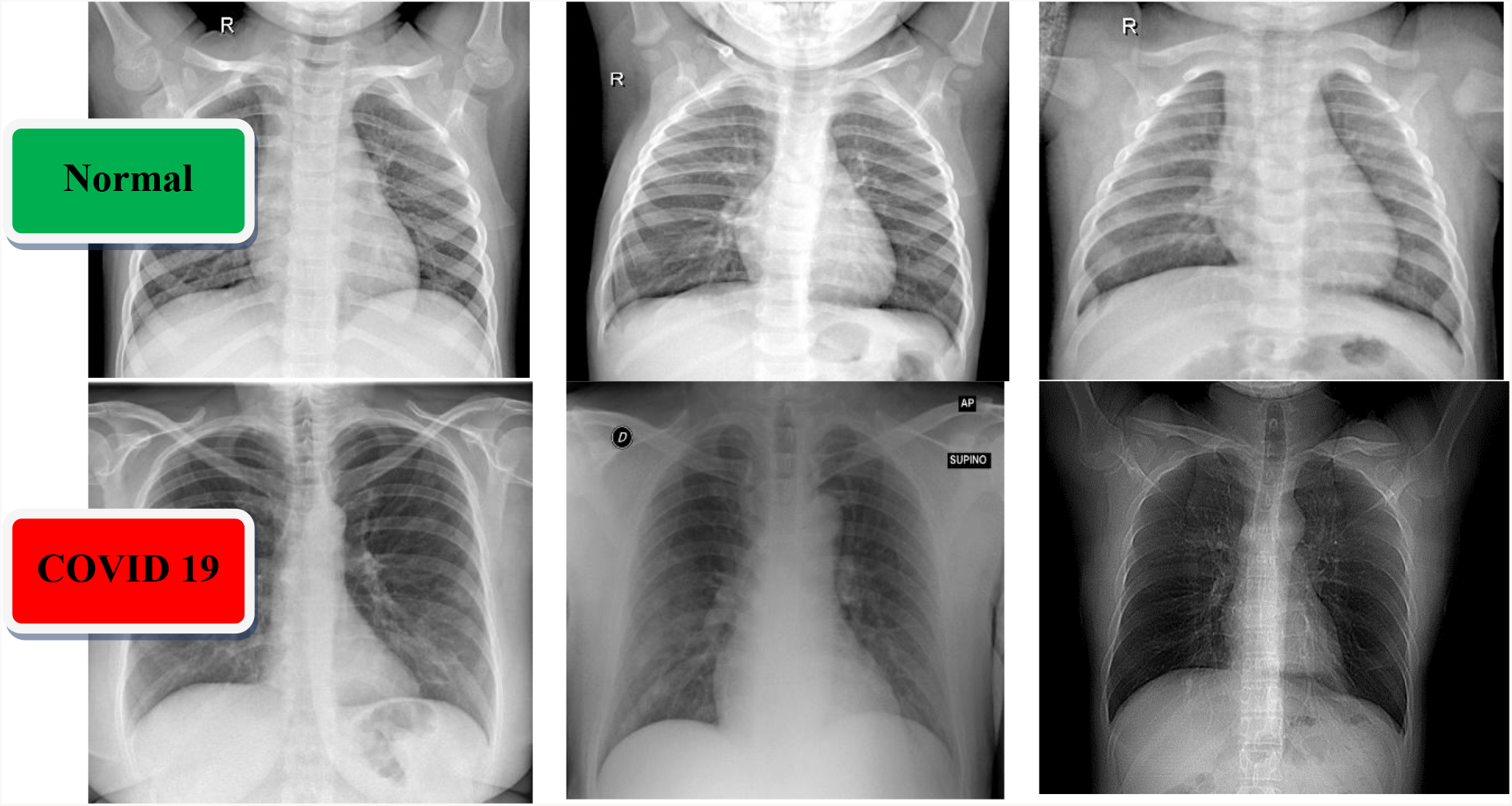
A sample of X-ray images dataset for normal cases (first row) and COVID-19 patients (second row).

### 2.2 Multi-Level Thresholding (MT)

A multi-level threshold is a process that splits grey image into several distinct areas. This technique specifies various image thresholds and splits the image into areas with certain intensities that correspond to one background and several objects [6]. The method works very well with coloured objects or complex backgrounds, cases in which a binary level threshold fails to realize satisfactory results is given in Eq. (1).

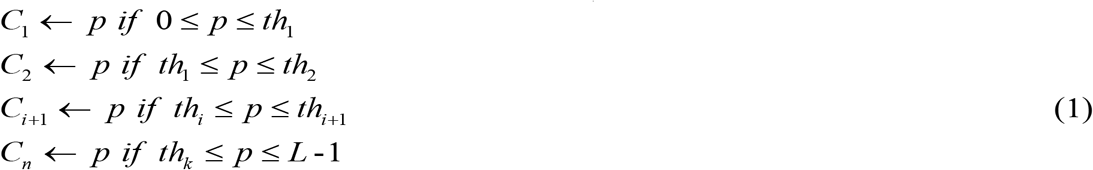

Where *p* is one of the pixels m × n that can be represented as one of the grayscale levels *L*1= {0, 1, 2, ‥, *L*-1}, C_1_ and C_2_ are classes with pixels p, representing {th_1_, th_2_, …, th_i_, th_i + 1_, th_k_} different thresholds.

### 2.3 Support Vector Machine (SVM)

SVMs are supervised learning algorithms that can be conducted for both classification or regression applications [22, 23]. SVM optimizes hyperplanes distance and the margin. Hyperplane distance can be maximized based on two-class boundaries using the following equations [24, 25].

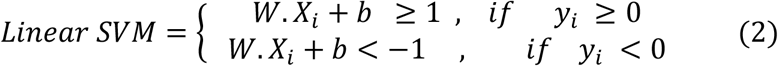

For i = 1,2,3, …… m. a positive slack variable (*ξ*) is added for handling the non-linearity as displayed in equation (3).

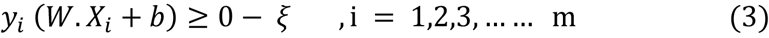

Accordingly, the objective function will be as an equation (4).

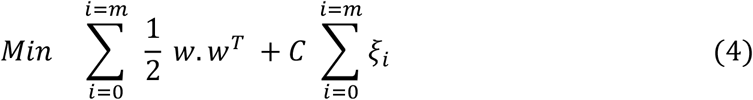

Where *W* is the weights matrix, *X* is the input vector, *b* is the bias vector, *y* is the output classes, *ξ*_*i*_ is the slack variable.

## 3. The Proposed Automatic COVID-19 lung classification System

The proposed system starts by visualizing a patient’s X-ray lung image and applying a median filter to enhance the contrast of the input images. Then, a multi-level image segmentation threshold based on Otsu objective function is applied. Then, the support vector machine has applied to classify the infected lung from non-infected. The overall procedures are described in detail in the following section along with the steps for each process. The overall architecture of the proposed COVID-19 classification system is described in Figure 2.

**Figure 2.**
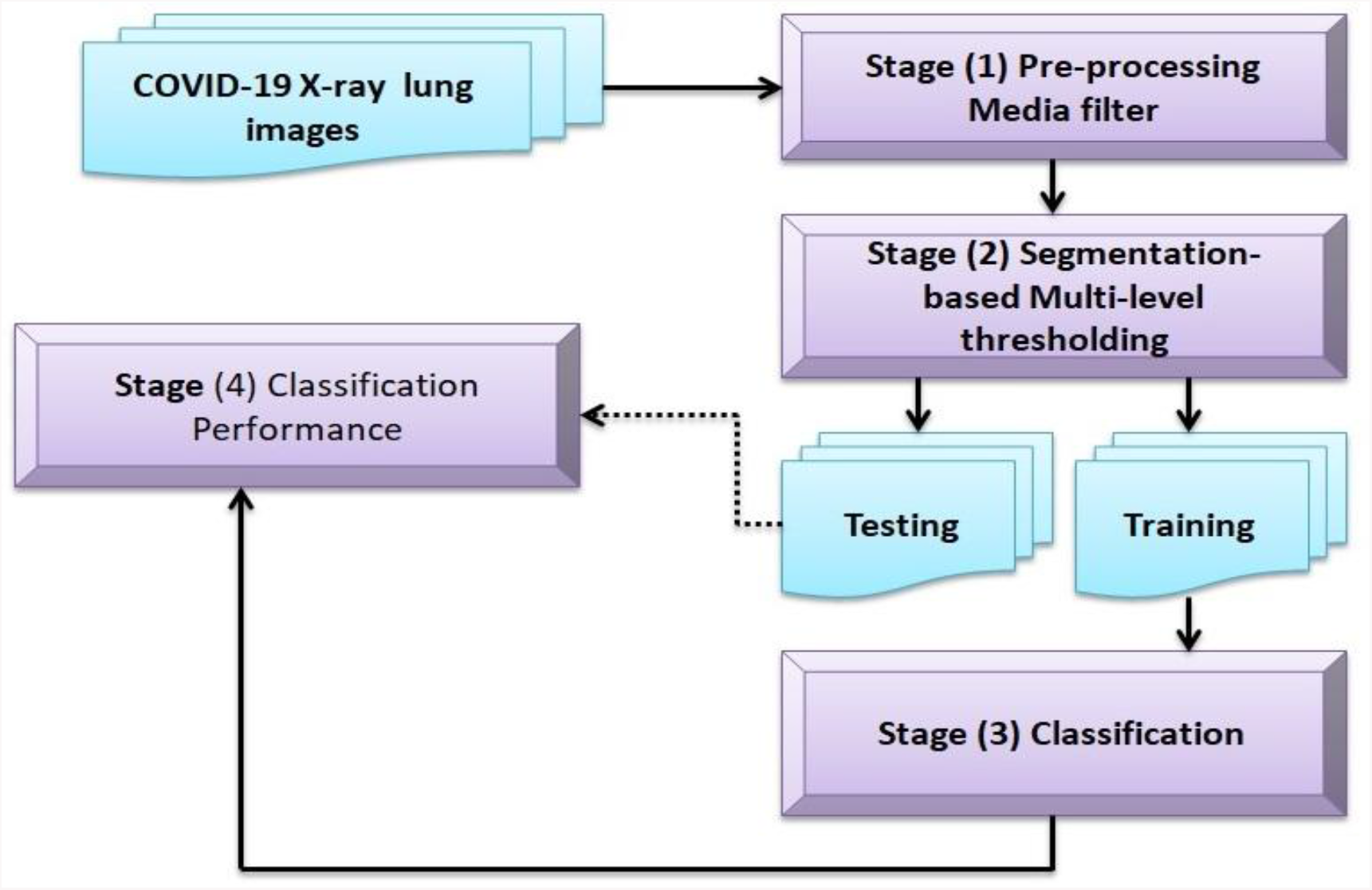
The overall COVID-19 classification system

**Figure 3.**
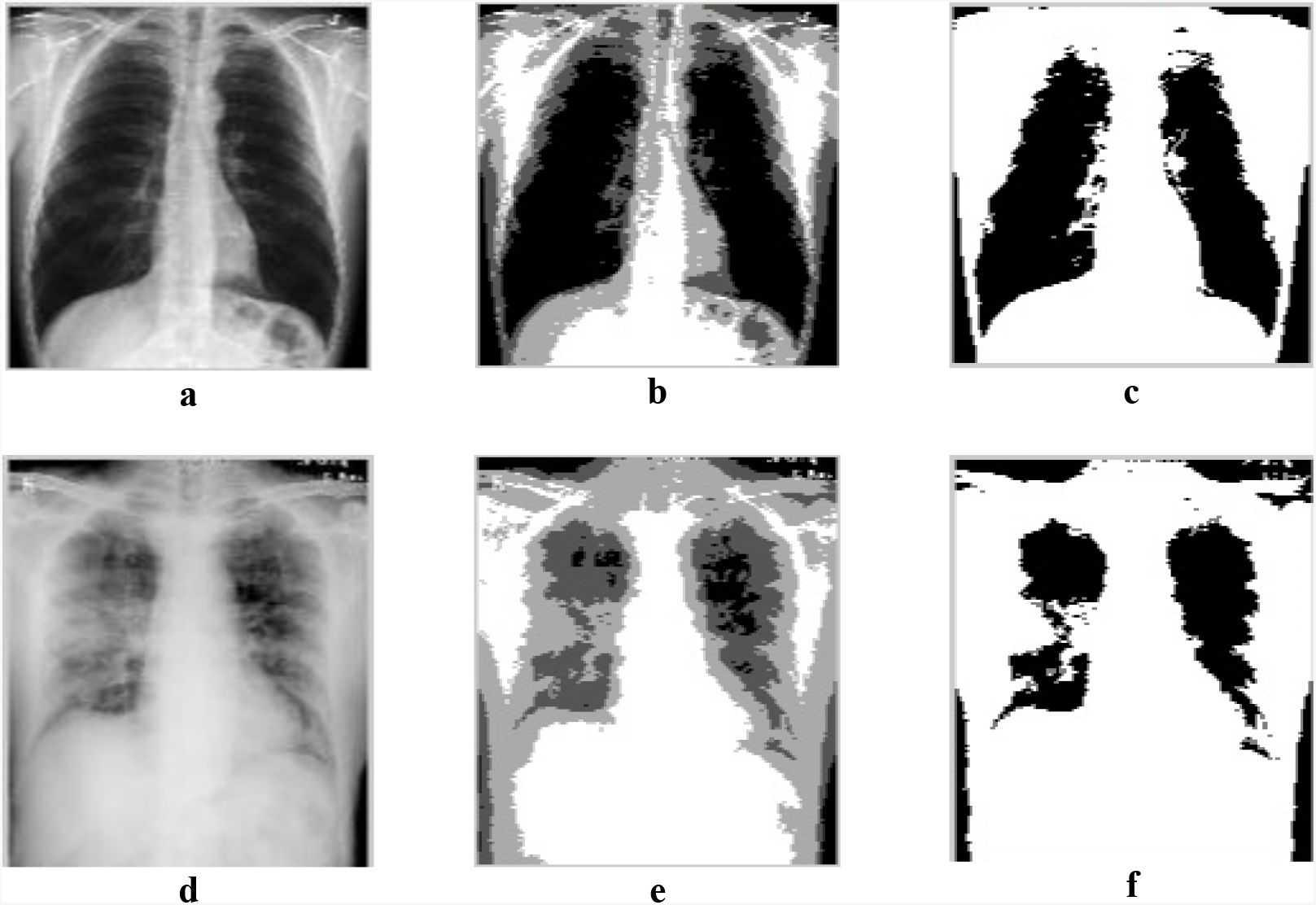
(a) image of normal case with normal lungs, (b) multi-level effects on the normal lung image, (c) final normal lung image depend on SVM respectively, (d) image of Covid-19 case with infected lungs, (e) multi-level effects on the infected lung image,(f) final infected lung image depend on SVM.

## 4. Experimental Results and Discussion

This study examined the performance of classification models for identification COVID-19. The experimental studies were implemented using the MATLAB 2019a deep learning toolbox. The results were obtained using a laptop equipped with an Intel Core i7, 18 GB of RAM and an AMD Radeon GPU. MATLAB was used to execute all the graphics and visualization functions. This model reads the data from a graphics file in the MATLAB workspace. Then multi-level thresholding was conducted to reduce number of objects in lung image the supported vector machine was applied to classify infected lung with COVID-19. Figure 4 (a-c) shows the image of normal case with normal lungs, Multi-level effects on the normal lung image and the final normal lung image depends on SVM respectively. Figure 4 (d-f) shows the image of COVID-19 case with infected lungs, Multi-level effects on the infected lung image and the final infected lung image depend on SVM, respectively. Sensitivity, specificity, accuracy, for the proposed system. The average sensitivity, specificity, and accuracy of the lung classification using the proposed model results were 95.76%, 99.7%, and 97.48%, respectively as illustrated in Table.1.

**Table.1.** Sensitivity, specificity, accuracy, for the proposed system

| Parameter | TPR (%) | FNR (%) | TNR (%) | FPR (%) | Sensitivity (%) | Specificity (%) | Accuracy (%) |
| --- | --- | --- | --- | --- | --- | --- | --- |
| **Case No** |  |  |  |  |  |  |  |
| **1** | 95.37 | 4.63 | 99.85 | 0.15 | 95.37 | 99.85 | 97.61 |
| **2** | 97.65 | 2.35 | 99.98 | 0.02 | 97.65 | 99.98 | 98.815 |
| **3** | 95.7 | 5.3 | 99.8 | 0.2 | 95.7 | 99.8 | 97.75 |
| **4** | 94.36 | 5.64 | 99.67 | 0.33 | 94.36 | 99.67 | 97.015 |
| **5** | 93.3 | 6.7 | 99.2 | 0.8 | 93.3 | 99.2 | 96.25 |
| **Average** | 95.276 | 4.924 | 99.7 | 0.3 | 95.276 | 99.7 | 97.48 |

## 5. Conclusion and future work

In the end of March 2020, more than +724000 confirmed cases of COVID 19 and more than +34000 deaths are exist globally where the humanity in our plant currently lives in COVID 19 pandemic. Isolation and social distance are temporary unpractical solution against fighting COVID-19. Unfortunately, a coronavirus vaccine is expected to take at least 18 months if it works at all. Moreover, COVID-19 pandemics can mutate into a more aggressive form [8]. Therefore, this paper presents a novel COVID-19 detecting methodology based on multi-level thresholding and SVM for X-ray images. The technique is useful for the clinical practitioner for early detection of COVID-19 infected patient. The model presents high accuracy where the average sensitivity, specificity, and accuracy of the lung classification were 95.76%, 99.7%, and 97.48%, respectively. Machine learning algorithms can present high performance in terms of accuracy and computational complexity [26]. Therefore, the future research may be based on using a modified model of optimized SVM or hybrid ML models. Moreover, larger data set can be providing higher model generalization.

## Data Availability

https://github.com/ieee8023/covid-chestxray-dataset

## References

[1] https://github.com/ieee8023/covid-chestxray-dataset].

[2] Nishiura, H., Linton, N. M., & Akhmetzhanov, A. R. (2020). “Serial interval of novel coronavirus (COVID19) infections”. International Journal of Infectious Diseases. doi:10.1016/j.ijid.2020.02.060

[3] Perlman S, Netland J. “Coronaviruses post-SARS: update on replication and pathogenesis”. Nat. Rev. Microbiol. 2009 Jun;7(6):439–50.

[4] Chan JF, To KK, Tse H, Jin DY, Yuen KY. “Interspecies transmission and emergence of novel viruses: lessons from bats and birds”.Trends Microbiol. 2013 Oct;21(10):544–55.

[5] Chen Y, Liu Q, Guo D. “Emerging coronaviruses: Genome structure, replication, and pathogenesis”. J. Med. Virol. 2020 Apr;92(4):418–423.

[6] Zhu, H.Q.; Xie, Q.Y. “A multiphase level set formulation for image segmentation using a MRF-based nonsymmetric Student’s-t mixture model”. Signal Image Video Process, 2018. vol.18, pp.1577–1585

[7] Jinkun Yang, Zhong Chen, Jiahao Zhang, Changheng Zhang, Qianqian Zhou, and Jian Yang “HOG and SVM algorithm based on vehicle model recognition”, Proc. SPIE 11430, MIPPR 2019: Pattern Recognition and Computer Vision, 114300T (14 February 2020);

[8] H. H. Elmousalami and A. E. Hassanien, “Day Level Forecasting for Coronavirus Disease (COVID-19) Spread: Analysis, Modeling and Recommendations,” arXiv preprint 2003.07778, 2020.

[9] F. Hutter, L. Kotthoff, and J. Vanschoren, Automated Machine Learning: Springer, 2019.

[10] C. Molnar, Interpretable machine learning: Lulu. com, 2019.

[11] Allam, Zaheer, and David S. Jones. “On the Coronavirus (COVID-19) Outbreak and the Smart City Network: Universal Data Sharing Standards Coupled with Artificial Intelligence (AI) to Benefit Urban Health Monitoring and Management.” Healthcare. Vol. 8. No. 1. Multidisciplinary Digital Publishing Institute, 2020.

[12] Novel, Coronavirus Pneumonia Emergency Response Epidemiology. “The epidemiological characteristics of an outbreak of 2019 novel coronavirus diseases (COVID-19) in China.” Zhonghua liu xing bing xue za zhi= Zhonghua liuxingbingxue zazhi 41.2 (2020): 145.

[13] Pan, Feng, et al. “Time course of lung changes on chest CT during recovery from 2019 novel coronavirus (COVID-19) pneumonia.” Radiology (2020): 200370.

[14] Chen, Huijun, et al. “Clinical characteristics and intrauterine vertical transmission potential of COVID-19 infection in nine pregnant women: a retrospective review of medical records.” The Lancet 395.10226 (2020): 809–815.

[15] Lim, Jaegyun, et al. “Case of the index patient who caused tertiary transmission of COVID-19 infection in Korea: the application of lopinavir/ritonavir for the treatment of COVID-19 infected pneumonia monitored by quantitative RT-PCR.” Journal of Korean medical science 35.6 (2020).

[16] Fang, Yicheng, et al. “Sensitivity of chest CT for COVID-19: comparison to RT-PCR.” Radiology (2020): 200432.

[17] Lan, Lan, et al. “Positive RT-PCR test results in patients recovered from COVID-19.” Jama (2020).

[18] Shuai Wang, Bo Kang, Jinlu Ma, Xianjun Zeng, Mingming Xiao, Jia Guo, Mengjiao Cai, Jingyi Yang, Yaodong Li, Xiangfei Meng, et al. A deep learning algorithm using CT images to screen for corona virus disease (COVID-19). medRxiv preprint medRxiv:2020.02.14.20023028, 2020a.

[19] Jun Chen, Lianlian Wu, Jun Zhang, Liang Zhang, Dexin Gong, Yilin Zhao, Shan Hu, Yonggui Wang, Xiao Hu, Biqing Zheng, et al. Deep learning-based model for detecting 2019 novel coronaviruspneumonia on high-resolution computed tomography: a prospective study. medRxiv preprintmedRxiv:2020.02.25.20021568, 2020b.

[20] Xiaowei Xu, Xiangao Jiang, Chunlian Ma, Peng Du, Xukun Li, Shuangzhi Lv, Liang Yu, Yanfei Chen, Junwei Su, Guanjing Lang, et al. Deep learning system to screen coronavirus disease 2019pneumonia. arXiv preprint 2002.09334, 2020.

[21] Ying Song, Shuangjia Zheng, Liang Li, Xiang Zhang, Xiaodong Zhang, Ziwang Huang, Jianwen Chen, Huiying Zhao, Yusheng Jie, Ruixuan Wang, et al. Deep learning enables accurate diagnosis of novel coronavirus (COVID-19) with CT images. medRxiv preprint medRxiv:2020.02.23.20026930, 2020.

[22] C. J. Burges, “A tutorial on support vector machines for pattern recognition,” Data mining and knowledge discovery, vol. 2, pp. 121–167, 1998.

[23] V. Vapnik, Estimation of dependences based on empirical data: Springer Science & Business Media, 2006.

[24] Elmousalami HH. Artificial Intelligence and Parametric Construction Cost Estimate Modeling: State-of-the-Art Review. Journal of Construction Engineering and Management. 2020 Jan 1;146(1):03119008.

[25] P. Mantero, G. Moser, and S. B. Serpico, “Partially supervised classification of remote sensing images through SVM-based probability density estimation,” IEEE Transactions on Geoscience and Remote Sensing, vol. 43, pp. 559–570, 2005.

[26] Elmousalami HH. Comparison of Artificial Intelligence Techniques for Project Conceptual Cost Prediction: A Case Study and Comparative Analysis. IEEE Transactions on Engineering Management. 2020 Feb 24.

[27] M. Chung et al., “Pr es s Pr es,” 2019.

[28] J. F. Chan et al., “Articles A familial cluster of pneumonia associated with the 2019 novel coronavirus indicating person-to-person transmission?: a study of a family cluster,” Lancet, vol. 6736, no. 20, pp. 1–10, 2020.

